# The use and impact of community Care (Education) and Treatment Reviews (C(E)TRs) in people with intellectual disability and autistic people: a cohort and case-control study using electronic health records

**DOI:** 10.64898/2026.11.24.26368824

**Authors:** Brónagh McCoy, Afia Ali, Lauren Bell, Ben Carter, Kang Wang, Peizhe Yan, Huajie Jin, Freya Elise, Daniel Acton, Sujeet Jaydeokar, Nick Gore, Deborah Chinn, Angela Hassiotis, André Strydom, Hitesh Shetty, Johnny Downs, Robert Stewart, Rory Sheehan

**Affiliations:** Department of Forensic and Neurodevelopmental Sciences, Institute of Psychiatry, Psychology and Neuroscience, King’s College London, London, UK; Wolfson Institute of Population Health, Queen Mary University of London, London, UK; Department of Biostatistics & Health Informatics, Institute of Psychiatry, Psychology and Neuroscience, King’s College London, London, UK; Department of Health Service & Population Research, Institute of Psychiatry, Psychology and Neuroscience, King’s College London, London, UK; NIHR Policy Research Unit: Mental Health, University College London, London, UK; Cheshire and Wirral Partnership NHS Foundation Trust – Centre for Autism Neurodevelopmental Disorders and Intellectual Disability (CANDDID), Chester, UK; Intellectual Disabilities Research Institute (IDRIS), University of Birmingham, Birmingham, UK; Faculty of Nursing, Midwifery and Palliative Care, King’s College London, London, UK; Division of Psychiatry, University College London, London, UK; NIHR Maudsley Biomedical Research Centre, London, UK; Department of Child & Adolescent Psychiatry, Institute of Psychiatry, Psychology and Neuroscience, King’s College London, London, UK and South London and Maudsley NHS Foundation Trust, London, UK; Department of Psychological Medicine, Institute of Psychiatry, Psychology and Neuroscience, King’s College London, London, United Kingdom and South London and Maudsley NHS Foundation Trust, London, UK

## Abstract

**Background:** Care (Education) and Treatment Reviews (C(E)TRs) were introduced in 2015 to prevent unnecessary hospital admissions amongst people with intellectual disability and autistic people. Little is known about people who receive community C(E)TRs or the effectiveness of community C(E)TRs in reducing admissions.

**Aims:** To estimate the causal effect of community C(E)TRs on psychiatric hospital admission, and compare length of hospital stay between patients who did and did not have a pre-admission community C(E)TR.

**Method:** We conducted a retrospective observational study using de-identified electronic health record data from 2,495 patients. We characterised the sociodemographic and clinical profiles of people with intellectual disability and autistic people who received a community C(E)TR between 2015 and 2025. We used propensity score weighting and a difference–in–differences analysis to compare changes in psychiatric hospital admissions in the six months before and after community C(E)TRs, with changes over corresponding periods in matched controls. Kaplan–Meier analysis was used to compare lengths of hospital stay between patients who had a pre–admission C(E)TR and those who did not.

**Results:** 446 community C(E)TRs were identified in the intellectual disability group and 382 in the autistic group. The intellectual disability group had a median age of 21 years and were predominately male (60.7%) whereas the autistic group were younger (median age 16 years) with a higher proportion of females (55.2%). Community C(E)TRs reduced the odds of psychiatric hospital admission over six months compared with controls (pooled OR = 0.47, 95% CI 0.26–0.85) but there was no difference over 12 months (pooled OR = 0.81, 95% CI 0.48–1.35). Length of hospital stay did not differ between those with and without a pre–admission C(E)TR.

**Conclusions:** These findings suggest that C(E)TRs may delay rather than prevent admissions and highlight the need for adequately resourced, sustainable community services to support people at high risk of admission.

## Introduction

Intellectual disability and autism are lifelong neurodevelopmental conditions, each present in 1–2% of the population. People with intellectual disability and autistic people experience higher rates of mental disorder than those without these conditions, reflecting a combination of biological, psychological, and social vulnerabilities^1,2^. A significant minority of people with intellectual disability and autistic people display behaviours that challenge, which may contribute to psychiatric hospital admission when associated with significant risks^3,4^.

Although psychiatric hospital admission is sometimes necessary, there have been longstanding concerns about an over-reliance on in-patient care for people with intellectual disability and autistic people, both in England and internationally^5–9^. These concerns include prolonged admissions without clear therapeutic goals, the use of restrictive practices such as physical restraint, and inappropriate psychotropic prescribing^10^.

Care (Education) and Treatment Reviews (C(E)TRs) were introduced through national policy published in 2015 as structured person-centred review meetings ^5^, chaired by a commissioner, which involve two independent advisors (a clinical expert and an expert-by-experience), the person, their family or carers, and relevant professionals. A community C(E)TR is convened when someone is considered at high risk of psychiatric hospital admission, with the aim of identifying and mobilising community supports that could enable the person to remain living safely in the community. If admission is required, a community C(E)TR can define therapeutic goals and establish a clear pathway towards discharge.

There has been very limited evaluation of community C(E)TRs, including who receives them and whether they reduce psychiatric hospital admissions or length of stay. A small clinical audit found that few admitted patients had a pre-admission C(E)TR and those who had experienced longer hospital stays than those who did not^11^. A time-series analysis using national data found that higher rates of pre-admission C(E)TRs were associated with a higher ratio of psychiatric hospital admissions to discharges^12^. These findings may reflect greater clinical complexity and higher admission risk of people receiving community C(E)TRs, rather than effects of the reviews themselves. However, as the first study included only admitted patients and the second used aggregated inpatient data, neither could establish whether community C(E)TRs prevent hospital admission.

In this study, we aimed to 1) characterise the sociodemographic and clinical profiles of people receiving community C(E)TRs, 2) estimate the effect of community C(E)TRs on psychiatric hospital admissions, and 3) examine the association between community C(E)TRs and length of hospital stay amongst those admitted.

## Method

### Study design and population

The protocol for this retrospective observational study was published before data analysis^13^. Data were extracted from the South London and Maudsley (SLaM) NHS Foundation Trust electronic health record (EHR) using the Clinical Record Interactive Search (CRIS) system. The study included people with a recorded diagnosis of intellectual disability or autism, ascertained from structured fields (ICD-10 diagnostic codes F70–79 and any F84.*, respectively) and through free text searches across individuals’ EHR using natural language processing (NLP)^14^ (also see Supplementary Text S1).

Descriptive analyses were conducted separately for people with intellectual disability with or without co-occurring autism, and autistic people without intellectual disability. Diagnostic groups were combined for the difference-in-differences analysis to increase statistical power.

### Procedures

Eligible participants had at least a one-year continuous period of care within a SLaM community mental health service between January 2015 and December 2023. C(E)TRs were identified in the EHR using free-text searches (Supplementary Text S2), with dates cross-checked against a record of in-patient admissions to ensure that only community C(E)TRs were included. Relevant demographic and clinical data (e.g. year of birth, diagnoses) were extracted.

Up to five controls (people who did not have a community C(E)TR) were extracted for each case (those with a community C(E)TR) based on age (±3 years), sex, and total care episode length before the date of the community C(E)TR (index date, ±2 years). Controls were assigned a pseudo-index date corresponding to the C(E)TR date. Controls needed to have an active community team episode and no inpatient episode on their assigned pseudo–index date. Individual controls could be matched to multiple C(E)TR events. A preprocessing pipeline was applied to ensure distinct community C(E)TRs were included with a requirement for active SLaM involvement in the six-month periods before and after each C(E)TR (Figure 1 and Supplementary Text S3).

**Fig 1.**
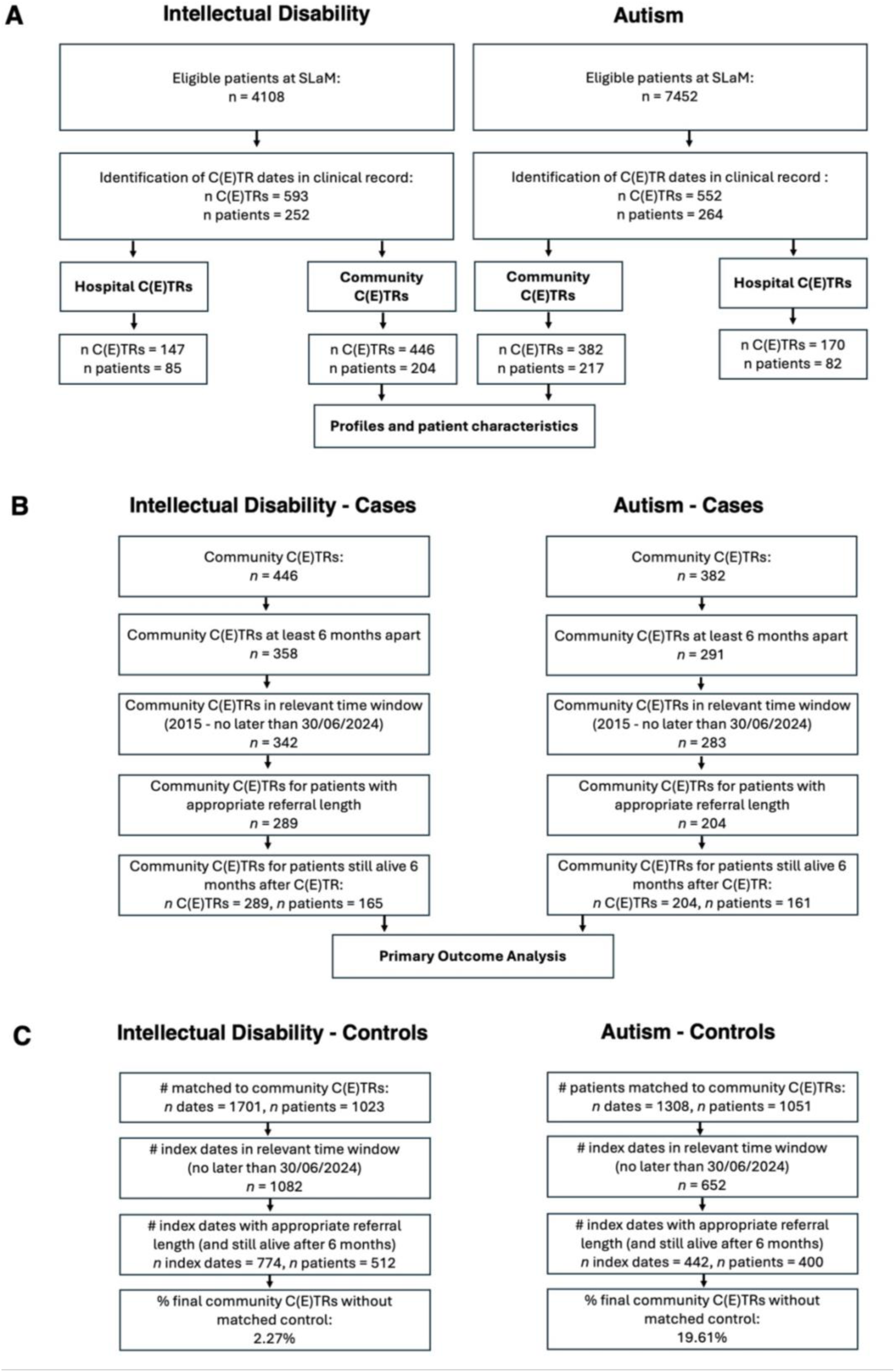
Flowchart. (A) Identification of all C(E)TRs and extraction of community C(E)TRs. Profiles and patient characteristics are presented using these data. (B) Cases: Pre-processing was applied to the community C(E)TRs in (A) to remove cases not meeting the inclusion criteria. The primary outcome analysis was based on data emerging from this pipeline. (C) Controls: preprocessing of control data was applied to the controls that were initially matched to cases (on index date, age, sex, and total care episode length).

A separate dataset was extracted for all psychiatric hospital admissions to SLaM in-patient services lasting at least 14 days amongst people with intellectual disability or autistic people.

### Primary outcomes

Primary outcomes were psychiatric hospitalisation within 6 months of a community C(E)TR (or pseudo-index date for controls) and length of psychiatric hospital stay amongst patients who were admitted.

### Descriptive and secondary outcome variables

Variables included age, sex, ethnicity, socioeconomic deprivation (measured using the Index of Multiple Deprivation (IMD) 2019), psychiatric diagnoses (by ICD–10 sub-chapter), psychotropic medication prescription (antipsychotic, mood stabiliser, anti-depressant, hypnotic/anxiolytic, medication for ADHD) (see Table 1). We compared the six–month periods before and after each community C(E)TR (or pseudo-index date for controls) for recorded psychiatric symptoms (e.g. aggression, agitation, suicidal thoughts), number of face–to–face community contacts with mental health professionals, and recorded mentions of Home Treatment Team (HTT), care package, or home care interventions ascertained using relevant NLP applications. A clinician–rated outcome measure, the Health of the Nation Outcome Scales (HoNOS)^15^, was also extracted within six months before or after the community C(E)TR, where available (Supplementary Text S4).

**Table 1.**
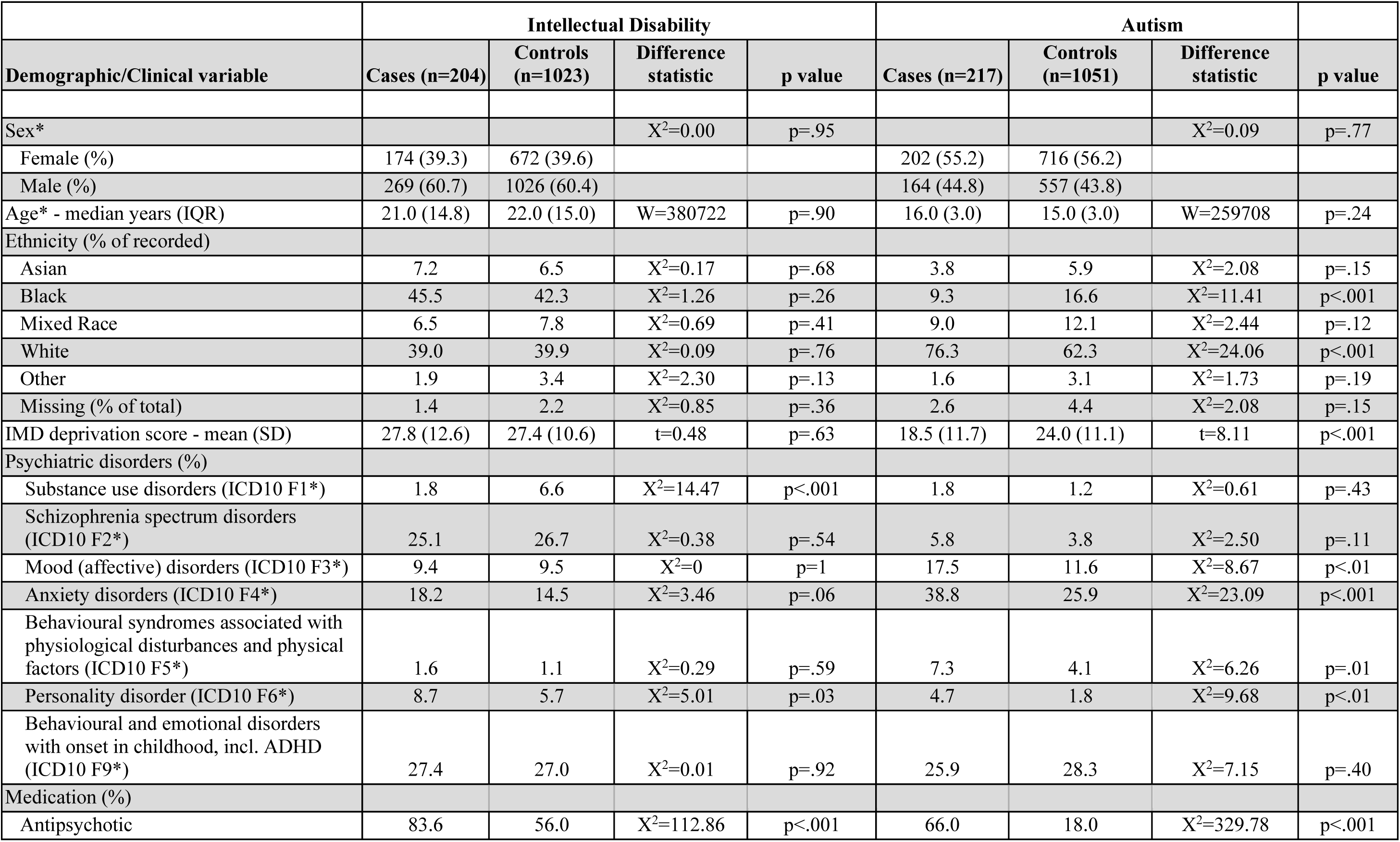
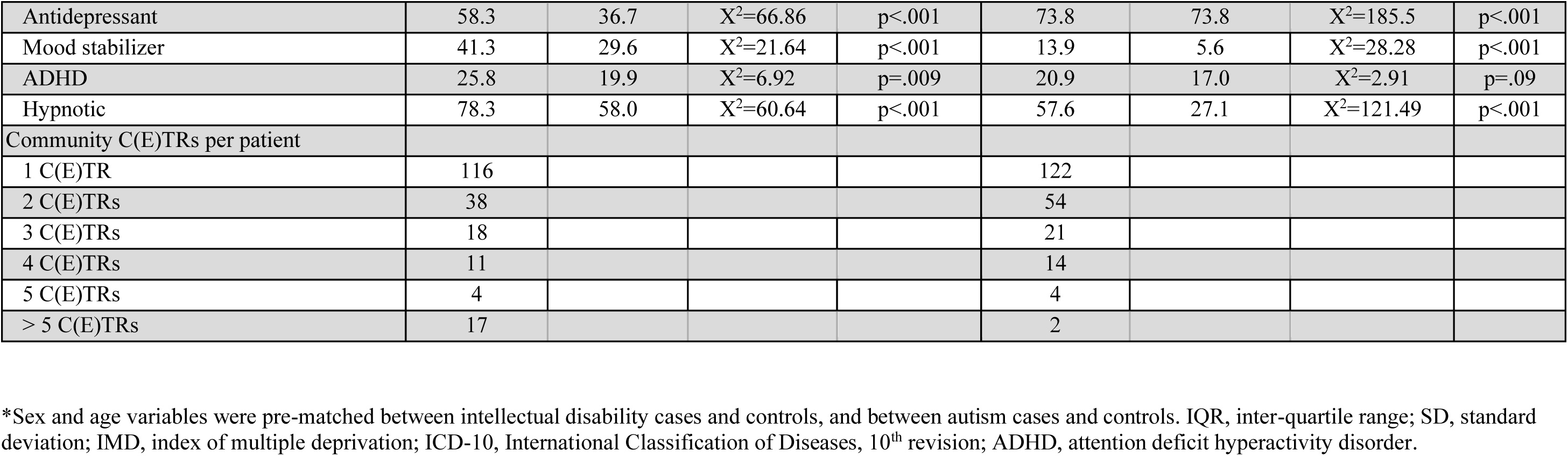
Characteristics of people receiving community C(E)TRs.

|  | Intellectual Disability |  |  |  | Autism |  |  |  |
|---|---|---|---|---|---|---|---|---|
| Demographic/Clinical variable | Cases (n=204) | Controls (n=1023) | Difference statistic | p value | Cases (n=217) | Controls (n=1051) | Difference statistic | p value |
| Sex* |  |  | X<sup>2</sup>=0.00 | p=.95 |  |  | X<sup>2</sup>=0.09 | p=.77 |
| Female (%) | 174 (39.3) | 672 (39.6) |  |  | 202 (55.2) | 716 (56.2) |  |  |
| Male (%) | 269 (60.7) | 1026 (60.4) |  |  | 164 (44.8) | 557 (43.8) |  |  |
| Age* - median years (IQR) | 21.0 (14.8) | 22.0 (15.0) | W=380722 | p=.90 | 16.0 (3.0) | 15.0 (3.0) | W=259708 | p=.24 |
| Ethnicity (% of recorded) |  |  |  |  |  |  |  |  |
| Asian | 7.2 | 6.5 | X<sup>2</sup>=0.17 | p=.68 | 3.8 | 5.9 | X<sup>2</sup>=2.08 | p=.15 |
| Black | 45.5 | 42.3 | X<sup>2</sup>=1.26 | p=.26 | 9.3 | 16.6 | X<sup>2</sup>=11.41 | p<.001 |
| Mixed Race | 6.5 | 7.8 | X<sup>2</sup>=0.69 | p=.41 | 9.0 | 12.1 | X<sup>2</sup>=2.44 | p=.12 |
| White | 39.0 | 39.9 | X<sup>2</sup>=0.09 | p=.76 | 76.3 | 62.3 | X<sup>2</sup>=24.06 | p<.001 |
| Other | 1.9 | 3.4 | X<sup>2</sup>=2.30 | p=.13 | 1.6 | 3.1 | X<sup>2</sup>=1.73 | p=.19 |
| Missing (% of total) | 1.4 | 2.2 | X<sup>2</sup>=0.85 | p=.36 | 2.6 | 4.4 | X<sup>2</sup>=2.08 | p=.15 |
| IMD deprivation score - mean (SD) | 27.8 (12.6) | 27.4 (10.6) | t=0.48 | p=.63 | 18.5 (11.7) | 24.0 (11.1) | t=8.11 | p<.001 |
| Psychiatric disorders (%) |  |  |  |  |  |  |  |  |
| Substance use disorders (ICD10 F1*) | 1.8 | 6.6 | X<sup>2</sup>=14.47 | p<.001 | 1.8 | 1.2 | X<sup>2</sup>=0.61 | p=.43 |
| Schizophrenia spectrum disorders (ICD10 F2*) | 25.1 | 26.7 | X<sup>2</sup>=0.38 | p=.54 | 5.8 | 3.8 | X<sup>2</sup>=2.50 | p=.11 |
| Mood (affective) disorders (ICD10 F3*) | 9.4 | 9.5 | X<sup>2</sup>=0 | p=1 | 17.5 | 11.6 | X<sup>2</sup>=8.67 | p<.01 |
| Anxiety disorders (ICD10 F4*) | 18.2 | 14.5 | X<sup>2</sup>=3.46 | p=.06 | 38.8 | 25.9 | X<sup>2</sup>=23.09 | p<.001 |
| Behavioural syndromes associated with physiological disturbances and physical factors (ICD10 F5*) | 1.6 | 1.1 | X<sup>2</sup>=0.29 | p=.59 | 7.3 | 4.1 | X<sup>2</sup>=6.26 | p=.01 |
| Personality disorder (ICD10 F6*) | 8.7 | 5.7 | X<sup>2</sup>=5.01 | p=.03 | 4.7 | 1.8 | X<sup>2</sup>=9.68 | p<.01 |
| Behavioural and emotional disorders with onset in childhood, incl. ADHD (ICD10 F9*) | 27.4 | 27.0 | X<sup>2</sup>=0.01 | p=.92 | 25.9 | 28.3 | X<sup>2</sup>=7.15 | p=.40 |
| Medication (%) |  |  |  |  |  |  |  |  |
| Antipsychotic | 83.6 | 56.0 | X<sup>2</sup>=112.86 | p<.001 | 66.0 | 18.0 | X<sup>2</sup>=329.78 | p<.001 |
| Antidepressant | 58.3 | 36.7 | X<sup>2</sup>=66.86 | p<.001 | 73.8 | 73.8 | X<sup>2</sup>=185.5 | p<.001 |
| Mood stabilizer | 41.3 | 29.6 | X<sup>2</sup>=21.64 | p<.001 | 13.9 | 5.6 | X<sup>2</sup>=28.28 | p<.001 |
| ADHD | 25.8 | 19.9 | X<sup>2</sup>=6.92 | p=.009 | 20.9 | 17.0 | X<sup>2</sup>=2.91 | p=.09 |
| Hypnotic | 78.3 | 58.0 | X<sup>2</sup>=60.64 | p<.001 | 57.6 | 27.1 | X<sup>2</sup>=121.49 | p<.001 |
| Community C(E)TRs per patient |  |  |  |  |  |  |  |  |
| 1 C(E)TR | 116 |  |  |  | 122 |  |  |  |
| 2 C(E)TRs | 38 |  |  |  | 54 |  |  |  |
| 3 C(E)TRs | 18 |  |  |  | 21 |  |  |  |
| 4 C(E)TRs | 11 |  |  |  | 14 |  |  |  |
| 5 C(E)TRs | 4 |  |  |  | 4 |  |  |  |
| > 5 C(E)TRs | 17 |  |  |  | 2 |  |  |  |

\*Sex and age variables were pre-matched between intellectual disability cases and controls, and between autism cases and controls. IQR, inter-quartile range; SD, standard deviation; IMD, index of multiple deprivation; ICD-10, International Classification of Diseases, 10<sup>th</sup> revision; ADHD, attention deficit hyperactivity disorder.

### Statistical analysis

Data were analysed using R Studio version 4.4.0 and Stata version 18. Frequencies and percentages are reported for categorical variables and mean (SD) or median (IQR) for continuous and count variables according to their underlying distributions. Within-individual statistical differences in pre- vs. post-community C(E)TRs (or pre- vs. post-pseudo-index dates for controls) were captured using McNemar’s Chi-squared test, and between-group differences were calculated using the Wilcoxon rank sum test (Mann–Whitney U–test).

The primary outcome analysis used propensity weighting followed by difference–in–differences (DiD) modelling to estimate the effect of community C(E)TRs on psychiatric hospital admission. Propensity scores were calculated using a multivariable logistic regression model^16^ incorporating baseline (pre-C(E)TR / index date) variables considered to be potential confounders of the association between community C(E)TRs and hospital admission (included age, sex, ethnicity, deprivation, diagnostic group, psychiatric diagnosis, psychotropic medication prescription). Propensity scores were converted to weights via the inverse probability of treatment weighting method^17^. A DiD model^18^ was then used to estimate the effect of community C(E)TRs, by estimating the change in hospitalisation over time for the C(E)TR compared to control group, reported as an odds ratio (OR) (Supplementary Text S5). Propensity model variables were assessed for missingness, and multiple imputation by chained equations was applied to produce five complete datasets^19^. The primary outcome analysis was run separately to each of these datasets and results were merged using Rubin’s rules^20^.

Three sensitivity analyses were conducted on the odds of hospital admission: 1) excluding C(E)TRs where the exact date was uncertain, 2) extending the propensity score model to include recorded psychiatric symptoms and mental health service use in the six-month period before the C(E)TR / pseudo-index date; and 3) restricting the analysis to the first community C(E)TR for each individual.

Kaplan-Meier survival curves were plotted for length of psychiatric hospital stay for the admitted patients dataset and compared patients with and without a community C(E)TR in the six months preceding admission.

### Ethical approval

CRIS has overarching ethical approval from the Oxford Research Ethics Committee C (reference 23/SC/0257) and the current study was approved by the local CRIS Oversight Committee (reference 2455/24–088).

## Results

We identified 446 community C(E)TRs for 204 individuals with intellectual disability and matched 1,701 pseudo-index dates across 1,023 people with intellectual disability who acted as controls between 2015 and 2023 (Figure 1). There were 382 community C(E)TRs for 217 autistic people, matched to 1,308 pseudo-index dates across 1,051 autistic controls. The annual number of community C(E)TRs increased gradually and peaked in 2021, followed by a sharp decline, particularly in the autistic group (Supplementary Figure S1).

### Characteristics of people receiving a community C(E)TR

People with intellectual disability who received a community C(E)TR had a median age of 21 years (IQR 14.8) and 60.7% were male. Just under half were Black (45.5%), with the next most frequent ethnic group being White (39.0%) (Table 1). These characteristics are broadly consistent with those of the intellectual disability control group and the wider population of people with intellectual disability accessing SLaM services (Supplementary Text S6). Socioeconomic deprivation and the prevalence of psychiatric diagnoses were also similar between C(E)TR recipients and controls.

Autistic people who received a community C(E)TR had a median age of 16 years (IQR 3.0), with more females than males in the sample (55.2% female), in contrast to the wider SLaM autistic population, of whom approximately 65% are male (Supplementary Text S6). Compared with controls, a higher proportion of C(E)TR recipients were White (76.3% vs 62.3%; p<0.001) and a lower proportion were Black (9.3% vs. 26.6%; p<.001). Autistic people who received a community C(E)TR were significantly less deprived than controls (mean IMD 18.5 vs 24.0, p<.001) and had a higher prevalence of anxiety disorders (38.7% vs 25.9% of controls, *χ*^2^(1)=23.1, p<.001).

Psychotropic medication prescribing before the C(E)TR (or pseudo-index date) was substantially higher across all medication categories among both people with intellectual disability and autistic people who received a community C(E)TR compared with controls (Table 1).

### Impact of community C(E)TRs on six-month psychiatric hospitalisation

Hospitalisation data were combined and analysed across both intellectual disability and autism groups (289 community C(E)TRs for 165 patients in the intellectual disability group, and 204 C(E)TRs for 161 patients in the autistic group; Figure 1). In people with intellectual disability receiving a community C(E)TR, psychiatric hospital admissions decreased from 12.5% in the preceding six months to 8.7% in the subsequent six months, compared with an increase from 3.3% to 4.4% in the control group (Table 2). A similar reduction was observed in the autistic C(E)TR group of 13.2% in the preceding six months to 9.3% in the six months after the C(E)TR, relative to almost no change in controls (2·9% to 2·7%). A propensity score weighting (see Supplementary Table S1) and DiD analysis showed that community C(E)TRs led to a greater reduction in the odds of psychiatric hospital admission over six months compared with controls (OR = 0.47, 95% CI 0.26–0.85).

**Table 2.**
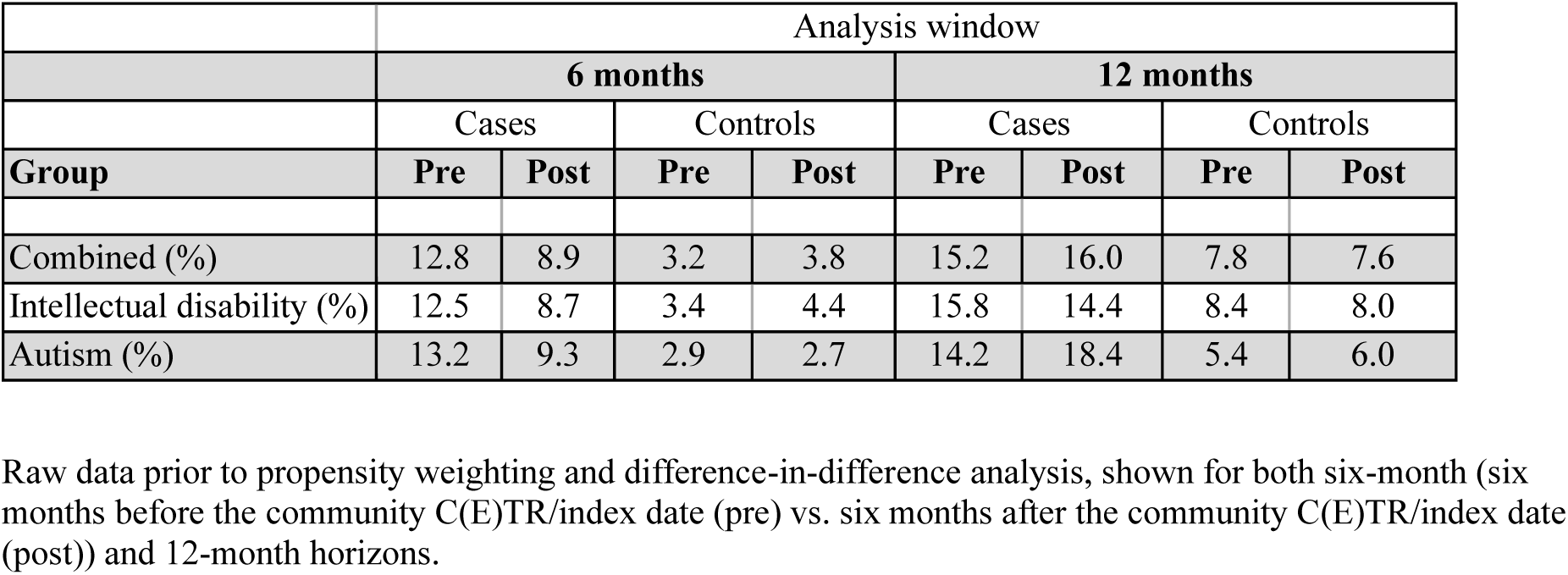
Psychiatric hospitalization rates.

|  | Analysis window |  |  |  |  |  |  |  |
|---|---|---|---|---|---|---|---|---|
|  | **6 months** |  |  |  | **12 months** |  |  |  |
|  | Cases |  | Controls |  | Cases |  | Controls |  |
| **Group** | **Pre** | **Post** | **Pre** | **Post** | **Pre** | **Post** | **Pre** | **Post** |
| Combined (%) | 12.8 | 8.9 | 3.2 | 3.8 | 15.2 | 16.0 | 7.8 | 7.6 |
| Intellectual disability (%) | 12.5 | 8.7 | 3.4 | 4.4 | 15.8 | 14.4 | 8.4 | 8.0 |
| Autism (%) | 13.2 | 9.3 | 2.9 | 2.7 | 14.2 | 18.4 | 5.4 | 6.0 |

Raw data prior to propensity weighting and difference-in-difference analysis, shown for both six-month (six months before the community C(E)TR/index date (pre) vs. six months after the community C(E)TR/index date (post)) and 12-month horizons.

### Sensitivity analyses

Excluding community C(E)TRs with uncertain dates (leaving 279 C(E)TRs in the intellectual disability group (753 matched control dates) and 185 C(E)TRs in the autistic group (411 matched control dates)) produced a similar result to the primary analysis (pooled OR = 0.48, 95% CI 0.20– 0.89). Extending the propensity score model to include additional clinical variables attenuated the effect of community C(E)TRs on psychiatric hospital admission (pooled OR = 0.64, 95% CI 0.34– 1.22). Restricting the analysis to the first recorded community C(E)TR date for each individual (165 C(E)TRs in the intellectual disability group (427 matched control dates) and 161 C(E)TRs in the autistic group (331 matched control dates)) also attenuated the effect (pooled OR = 0.90, 95% CI 0.43–1.91).

### Twelve-month impact of community C(E)TRs on psychiatric hospitalisation

The primary outcome analysis was extended to examine psychiatric hospital admissions over the 12 months before and after a community C(E)TR. After preprocessing, the analysis included 215 community C(E)TRs in 140 people with intellectual disability and 453 pseud–index dates in 311 controls, and 141 community C(E)TRs in 120 autistic people, matched to 168 pseudo-index dates in 161 controls. Hospitalisation proportions for the combined intellectual disability and autism cases were 15.2% in the 12 months before a C(E)TR and 16.0% in the 12 months after a C(E)TR (Table 2). The control groups had hospitalization proportions of 7.8% in the 12 months pre-index date and 7.6% in the 12 months after. After propensity weighting and DiD analysis, there was no clear evidence of an effect of community C(E)TRs on psychiatric hospital admission over 12 months compared with controls (pooled OR = 0.81, 95% CI 0.48–1.35).

### Changes in psychiatric symptoms and service use

Changes in recorded psychiatric symptoms and service use are shown in Supplementary Table S2. In people with intellectual disability receiving a community C(E)TR, the proportion of patients with a mention of aggression and agitation was significantly higher in the six-month period after compared to before a community C(E)TR, with no corresponding changes in controls (p<.05, see Supplementary Table S2). In autistic people who received a community C(E)TR, the proportion with a mention of suicidal ideation increased after a community C(E)TR, compared with a small reduction in controls (p<.05). Increases in recorded anxiety, aggression, agitation, and hostility were also significantly greater in autistic C(E)TR recipients than in controls (all p<.05). Too few patients had HoNOS measures in the relevant period for formal analysis (29 ID patients, Supplementary Text S4).

Mentions of care packages increased significantly following community C(E)TRs in both groups compared to controls (p<.05). There were no differences in the number of community contacts with healthcare professionals (Supplementary Table S3).

### Length of psychiatric hospital stay

A separate dataset was extracted for those patients who were admitted to psychiatric hospital. Of 545 admissions in the intellectual disability group (340 patients), only five admissions (five patients) were preceded by a community C(E)TR within six months. There were 308 admissions (202 patients) in the autism group, with ten of those admissions (ten patients) occurring within six months after a community C(E)TR. Across both groups, only 1.8% of psychiatric hospital admissions were preceded by a recent community C(E)TR. The small numbers precluded robust statistical comparisons of length of stay between groups. A Kaplan-Meier survival plot on combined data across both the intellectual disability and autism groups shows substantial overlap in admission length between admissions with and without a preceding community C(E)TR (Figure 2). Extending the period to 12 months before admission identified 9 admissions in the intellectual disability group and 17 in the autistic group with a preceding C(E)TR, again with no clear separation between the survival curves (Figure 2).

**Fig 2.**
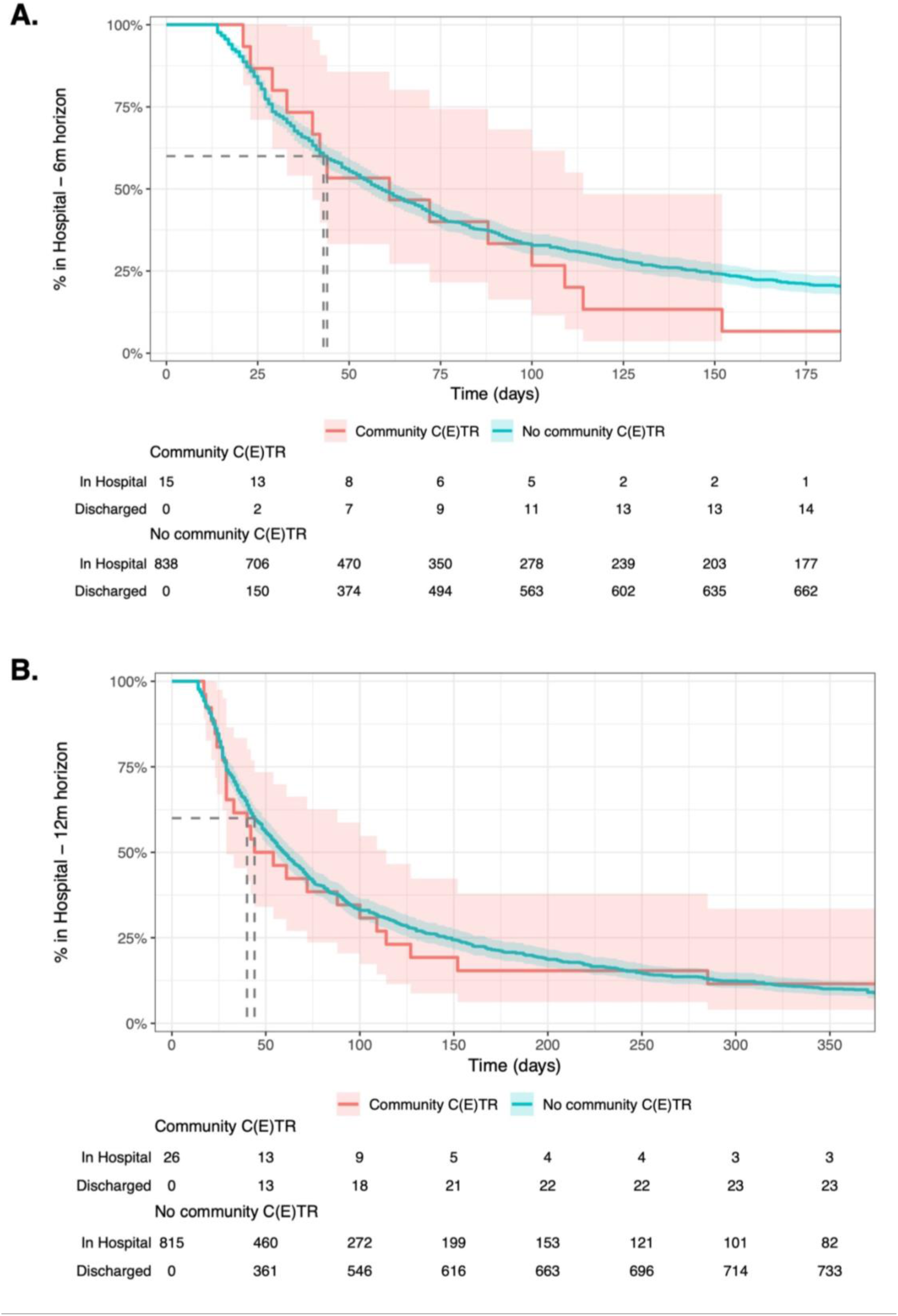
Kaplan-Meier survival curves. Length of stay for psychiatric hospital admissions of people with intellectual disability and/or autistic people. (A) Patients who had a community C(E)TR within 6 months prior to admission, compared to patients who did not. (B) patients who had a community C(E)TR within 12 months prior to admission, compared to patients who did not. Data is censored at 6 months (A) and 12 months (B) due to diminished size of C(E)TR group.

## Discussion

This study provides new evidence on the use and effectiveness of C(E)TRs, a cornerstone of national policy in England to improve support for people with intellectual disability and autistic people in the community and to prevent unnecessary hospital admissions. In our primary analysis, community C(E)TRs led to a 53% reduction in the odds of psychiatric hospital admission compared with controls. However, when we extended the analysis to admissions over a 12–month period, the reduction in admissions was no longer significant. This suggests that the benefits of a community C(E)TR may diminish over time; it may be that additional support is mobilised immediately following the C(E)TR, helping to avert an admission, which is then not sustained or sufficient to address the underlying factors contributing to risk of admission. Hence, in some cases, community C(E)TRs might delay rather than prevent admissions.

We found that community C(E)TRs were predominantly undertaken for adolescents and young adults. While the diagnostic and socio-demographic characteristics of people with intellectual disability who received community C(E)TRs was similar to their controls, autistic people who received a community C(E)TR had a lower level of deprivation compared to the control group and higher levels of anxiety disorders. The higher rates of anxiety disorders in autistic females, particularly in adolescents, is consistent with existing research in urban populations^21^. The young profile of C(E)TR recipients, particularly autistic people, may reflect differences in clinical need or historical under–recognition of autism in older adults^22^. Alternatively, the availability of keyworker support offered to younger people may facilitate the identification of unmet needs and coordination of C(E)TRs^23^. The higher proportion of White autistic people and a lower proportion of Black autistic people receiving community C(E)TRs compared to controls raise concerns about potential ethnic inequalities in access to reviews.

Across both diagnostic groups, C(E)TR recipients were more likely to be prescribed all categories of psychotropic medications compared to controls, despite recorded rates of psychiatric diagnoses being similar, which may reflect the presence of more severe or persistent symptoms in people who received a community C(E)TR, prescribing for behaviours of concern, or potentially limited access to non-pharmacological interventions.

The notable effect of community C(E)TRs on hospital admission was attenuated after additional adjustment for psychiatric symptoms and service use preceding the review. Although the point estimate remained below one, confidence intervals notably overlapped one. This suggests that differences in clinical presentation and service use between C(E)TR recipients and controls may partly explain the observed effect. Residual confounding by factors that are not adequately captured by routinely–collected data (e.g. severity, risk, and stability of community placements) may also be important. A sensitivity analysis taking only the first community C(E)TR substantially attenuated the six-month benefit in terms of reduction of admissions, although this was based on a smaller dataset and under-powered. It may be that repeated community C(E)TRs provide additional benefit perhaps through ongoing monitoring and follow-up of recommendations, although this requires further investigation.

We investigated whether community C(E)TR were associated with changes in clinical presentation or community support. Mentions of certain symptoms increased following community C(E)TRs (aggression and agitation in people with intellectual disability group, and suicidal ideation autistic people) which may reflect increased clinical scrutiny and documentation of concerns that prompted the review, rather than a deterioration in clinical status. There was no change in community contacts with mental health professionals following community C(E)TRs, although our data did not capture input from social care professionals or paid support staff. Mentions of a ‘care package’ increased following community C(E)TRs which potentially indicates the care and support that a person receives is formally reviewed or altered following a C(E)TR. However, the context of mentions in the patient record was not available, which limits interpretation of the data obtained through NLP methods.

Although some psychiatric hospital admissions are avoidable, inpatient care remains necessary when a person’s assessment or treatment needs cannot be safely met in the community^24^. In this case, a community C(E)TR may be beneficial to clarify the purpose of an admission and support early discharge planning, potentially reducing the length of stay. The small proportion of admissions that were preceded by a community C(E)TR in our study is consistent with national data^25^ and proved insufficient for a robust analysis on differences in length of stay between groups.

Our data show time trends in the delivery of community C(E)TRs, with the highest numbers seen during the COVID–19 period and immediately afterwards, followed by a marked decline. The increased use during the COVID–19 pandemic may reflect disruption to usual care and support, potentially increasing the risk of hospital admission. It is possible to deliver C(E)TRs remotely which may also have facilitated their delivery. The subsequent sharp decline is notable given the continued emphasis on C(E)TRs in national policy^26^. Possible explanations include greater reliance on less formal multi-disciplinary professional meetings or the Local Area Emergency Protocol (LAEP), and changing attitudes towards C(E)TRs amongst professionals, patients, and carers^27^. Convening a C(E)TR requires considerable resource and co-ordination across agencies which may discourage their use and the period covered by our study also includes significant changes to the organisation and commissioning of mental health services following publication of the NHS Community Mental Health Framework^28^.

### Clinical and research implications

Our findings demonstrate the potential value of community C(E)TRs in reducing admissions but also raise concerns about potential inequalities in access to reviews and declining implementation. Community C(E)TRs should form part of an integrated admission avoidance pathway, supported by effective use of local Dynamic Support Registers to identify people at risk of admission. Evidence-informed risk stratification and decision support tools could help improve consistency of decisions about when to convene a community C(E)TR, reducing reliance on subjective judgment^29,30^. Although community C(E)TRs may mobilise support in the short-term, panel recommendations are not binding and stronger mechanisms for monitoring implementation and ensuring accountability may improve their effectiveness. The low proportion of admissions preceded by a community C(E)TR raises the prospect of missed opportunities to define the purpose of admissions and plan for discharge from the outset. However, pre–admission reviews may not be feasible in emergency situations.

Future research is required to establish definitively the effectiveness of community C(E)TRs and their impact on a wider range of outcomes, including quality of life and restrictive practices, ideally using larger multisite datasets. Implementation research could explore barriers and facilitators to effective delivery and how community C(E)TRs interact within wider admission avoidance infrastructure, including LAEP meetings, Home Treatment Teams and Intensive Support Teams, in responding to escalating needs or crisis situations.

### Strengths and limitations

This study is the first large–scale evaluation of the use and impact of community C(E)TRs in routine clinical practice, using a systematic approach to identifying C(E)TRs from electronic health records. Longitudinal, individual-level data enabled us to examine characteristics of people receiving C(E)TRs as well as patterns of implementation over almost a decade, capturing changes in policy and practice over time. The use of quasi-experimental methods including propensity score weighting and difference–in–differences analysis strengthen our ability to make causal inferences about the effect of community C(E)TRs, and sensitivity analyses explored the robustness of our findings. Examining hospitalisation over six– and 12–month time horizons provided insights into potential short- and longer–term impacts of community C(E)TRs. The inclusion of all people with recorded intellectual disability or autism accessing SLaM services reduces recruitment and selection bias which may occur in studies in which people are recruited directly.

The relatively small number of community C(E)TRs limited statistical precision, particularly in the length of hospital stay analysis. Pooled estimates for the primary outcome analysis might obscure differences between the intellectual disability and autistic groups. Reliance on routinely–collected electronic health data has disadvantages, including some variables being missing or inadequately reported (e.g. HoNOS), and hospital admissions outside SLaM are not measured, although we expect them to be minimal. Residual confounding remains possible and the attenuation of the effect in sensitivity analyses demonstrates some uncertainty around the findings. We were unable to capture the full range of community services and interventions; this is particularly relevant for people with intellectual disability and autistic people who may receive substantial support from family or paid carers, social services, and third sector organisations. Our primary outcome was hospital admission and we could not determine whether admissions were potentially avoidable. Finally, the study was conducted in a single mental health Trust in London and findings may not be representative of other regions where the organisation of services and implementation of community C(E)TRs may differ.

## Conclusions

Community C(E)TRs may help to prevent hospital admissions for people with intellectual disability and autistic people in the short–term. However, their declining use and the small proportion of admissions preceded by a C(E)TR raises concerns about implementation. Achieving sustained reductions in unnecessary admissions will depend on equitable access to reviews and ensuring their recommendations are translated into meaningful and sustained changes in care and support.

## Contributors

RS, AA, BC, HJ, FE, DA, SJ, NG, DC, AH, AS, JD, and RSt contributed to study conceptualisation and funding acquired. BM and HS curated the data. BM conducted the formal analysis, administered the project, and created visualisations. BM, HS, KW, and PY contributed to the investigation. BM, LB, and BC developed the study methodology. RSt, HS, and JD provided CRIS resources. RS and AA supervised the work. BM, AA, and RS wrote the original draft. BM, AA, LB, BC, KW, HJ, FE, DA, SJ, NG, DC, AH, AS, JD, RSt, and RS reviewed and edited the manuscript. BM, HS, KW, and PY had full access to the data reported in the study. All authors contributed to interpretation of the findings, approved the final version of the manuscript, and accepted responsibility for the decision to submit for publication.

## Data availability

De-identified electronic health record data are not publicly available, and the data and corresponding analytic code may only accessed under SLaM CRIS governance. Researchers may apply for access through the CRIS oversight process, subject to approval and relevant information-governance requirements.

## Acknowledgments

We would like to acknowledge and thank the study’s patient and public involvement (PPI) groups for their contributions, especially Jessica Solly, Caroline Adlam, and Thomas Vaughan. We also thank Lesley Gledhill and Claire Evans for coordinating the PPI group of young people and adults with intellectual disability, and Freya Elise (co-author) as the lived experience representative and coordinator of the autistic adults PPI group. The authors made use of ChatGPT for the final draft of the manuscript to improve readability and language.

## Supplementary Appendix

### Supplementary Text S1. Extracting variables using Natural Language Processing (NLP) applications

NLP applications have been used to extract structured information from the free text of electronic health records at SLaM since 2009, and have been developed to target specific clinical symptoms, physical health conditions, and interventions/services mentioned in patient records. In this study we use NLP apps to identify diagnoses, medications, and free text mentions of psychiatric symptoms and interventions, such as e.g. aggression, for specific periods of time. For the ‘aggression’ app, for example, the associated search term was *agitat*, which included the past or present participle of the word. The development process for each app includes performance metrics on tested batches of documents for various populations. Information on the NLP apps for each symptom and intervention used in this study can be found in the Maudsley BRC CRIS NLP Applications Library^1^.

### Supplementary Text S2. CETR identification and reliability of CETR labelling

C(E)TRs were identified in the EHRs of the sample using the following key word search terms: “Care and Treatment Review”, “Care Education and Treatment Review”, “CTR”, or “CETR”. Records were reviewed to determine if and when C(E)TR meetings were held. Separate lists were made by two independent raters to reflect the level of confidence of a C(E)TR having occurred on a specific date. CETR identification and labelling were carried out by two independent raters. Of the pool of patients who had a learning disability diagnosis, 936 had a mention of one of the C(E)TR keyword search terms in their records The first 100 of these patients were used to pilot C(E)TR labelling and categorization. Patients were assigned to one of the five lists below, to capture the level of confidence in a C(E)TR having occurred on a specified date: 1) C(E)TR occurred on a specified date, 2) C(E)TR proposed for a specified date but no evidence it occurred, 3) C(E)TR previously occurred but without a specified date captured in the records, 4) C(E)TR mentioned/proposed for a patient with no specified date and no evidence it occurred, 5) False positive, with the search terms pulling back records that were not relevant to a C(E)TR occurring for that patient. Inter-rater reliability on the level of agreement of assigning patients to one of these five categories was calculated and indicated strong agreement between the two raters (Cohen’s k = 0·86). Three patient IDs were missed by one of the raters, so of the 97 remaining patients, 38 of these were confirmed to have had at least one C(E)TR occur on a specified date. Following up on those patients for whom raters agreed a C(E)TR was held, the percentage agreement on different aspects of these were as follows: agreement on date of one CETR: 92%; agreement on multiple CETRs having occurred: 89%, and agreement on dates of more than one CETR: 81%. The two raters then went through all records where there was disagreement to clarify misidentifications and/or alternative interpretations, to ensure their independent assessment of records of the remaining 836 patients (∼400 each) was fully aligned. A flag to denote the level of confidence in the date of the C(E)TR was also included (confident, not confident). The ‘not confident’ option mainly applied to two scenarios: 1) when the exact C(E)TR date was not clear but seemed have occurred between two records, and it was written about as having recently occurred in the second, reviewers include a note that refers to the date of the record where a CETR is mentioned as having recently happened, and 2) when a C(E)TR was mentioned as having occurred during a particular month, but without a date, we included the C(E)TR as having occurred on the 15^th^ of that month.

### Supplementary Text S3. Preprocessing for change in hospitalization primary outcome measure

We excluded 1) inpatient C(E)TRs, 2) C(E)TRs falling within 6 months of a previous community C(E)TR for the same patient, 3) C(E)TRs occurring after 30/06/2024 (to ensure full data captured within study time window), 4) patients with less than 6 months of data available following a C(E)TR due to e.g. being transferred to another service, and 5) patients who died within 6 months of having a C(E)TR (see figure 1 in manuscript for flowchart). As control patients were initially matched to cases based on sex, age, and referral length, we retained controls if they were a match with the remaining cases after preprocessing. Sociodemographic and clinical profiles of people undergoing community C(E)TRs were extracted before applying the rest of the preprocessing pipeline, i.e. anybody receiving a community C(E)TR, and their age- and sex-matched controls (manuscript Figure 1). The remaining preprocessing steps were applied before statistical analyses on psychiatric hospitalization and other clinical variables describing changes in the six months before versus six months after the index date.

### Supplementary Text S4. Health of the Nation Outcome Scales (HoNOS)

Data were extracted for C(E)TR and control patients for all versions of the HoNOS (HoNOS, HoNOS-LD and HoNOSCA, for standard, learning disability, and children and adolescents, respectively). For each patient, the closest HoNOS completed within 6 months before the C(E)TR was compared to the closest HoNOS within six months after the C(E)TR. The latter HoNOS must also have been administered at least six weeks after the C(E)TR, as anything captured within six weeks is unlikely reflective of changes to care emerging from the C(E)TR.

Few patients had eligible clinician-rated scales on psychiatric symptoms, social functioning, and overall wellbeing, captured by the HoNOS (see Methods). Data for the ID version of the scale, HoNOS-LD, was sufficient to carry out a preliminary analysis in the C(E)TR group only, with relevant pre- and post- C(E)TR scores for 44 ID cases (29 ID individuals). There was no difference in total HoNOS-LD scores after compared to before a community C(E)TR (pre: 17.83 ± 9.85 vs. post:16.95 ± 8.55; t(41)= –1.02, p=0.35).

### Supplementary Text S5. Causal framework for change in hospitalization primary outcome measure

The estimand for the difference-in-differences (DiD) model was the average treatment effect on the treated (ATT)^2^. The DiD method compared the six-month change in the psychiatric hospitalization outcome in patients who had a community C(E)TR to the change in the control group. The model was implemented as a logistic regression with psychiatric hospitalization (yes/no) as the outcome. We used the *xtlogit* function in Stata 18, suitable for panel data (within-subject repeated-measures), and applied population-averaging^3^ to capture the effect of community C(E)TRs at the group-level. The regression was set up according to the formula:

Yit = β1 * GROUP + β2 * PERIOD_it_ + β3 * GROUP * PERIOD_it_ + e_it_

Y is the outcome in psychiatric hospitalization (yes=1/no=0) of patient *i* at time *t*, GROUP is a dummy variable with a value of 1 for C(E)TR cases and 0 for controls, and PERIOD is a dummy variable with a value of 1 for the six-month period after a C(E)TR (or matched index date for controls). The coefficient of interest is β3, the coefficient on the interaction term GROUP * PERIOD referred to as the DiD parameter. It indicates the change in the outcome in the C(E)TR groups relative to the change in the outcome in the control groups. To balance the groups on important covariates, IPTW weights from the propensity model were applied to this DiD GROUP * PERIOD interaction term^2,4^.

#### Causal framework assumptions

The causal framework comes with several assumptions^5^. For DiD methods, the parallel trends assumption is that the treatment and control groups would have had the same trends across time if nobody had had a community C(E)TR. This assumption is not testable but was made more reasonable by the initial matching of the control group and propensity model IPTW adjustment for covariates. The two types of associated selection bias that can occur are: across time and across group. Our study design reduces concerns regarding selection bias across time because outcome measurements were obtained from the same individuals during both the pre- and post-periods. Thus, any observed differences were not driven by changes in the composition of either the C(E)TR or control groups over time.

We also ensured that we only included patients who were under SLaM services for the full six-months before and after the C(E)TR / index date. Furthermore, during initial matching, up to five controls were matched to a C(E)TR patient on age and sex, and to assign an index date to the controls that aligned with the C(E)TR date of the case. As such, pre- and post-index date time periods of controls were matched to their respective case pre- and post-C(E)TR dates, that could have occurred anytime between 2015 and 2025. Patients in both groups were therefore likely to be affected in a similar way by any other factors such as changes in clinical practice over time or the response at SLaM to the COVID pandemic. The initial propensity model was used to balance co-variates that may differ between treatment and control groups, thereby addressing the across-group selection bias that may undermine the parallel trends assumption.

The positivity assumption posits that all patients have a positive probability of being in each of the groups; here, all patients had a learning disability and/or were autistic, and were under secondary mental healthcare services at SLaM, and thus were all eligible to receive a community C(E)TR. Unconfoundedness assumes that assignment to the community C(E)TR group is not related to the hospitalization outcomes that would be observed in the controls, conditional on the observed covariates. The initial matching and propensity weighting model were established to match controls to C(E)TR patients on a range of (risk) variables to meet this assumption. We extracted all covariates from the period before (or on) the community C(E)TR date / index date, thereby meeting the exogeneity assumption of covariates not being affected by the treatment itself. Lastly, the stable unit treatment value assumption poses that each patient’s potential outcomes were not affected by the treatment assignments of any other patients; all patients included were independently receiving care, and each subsequent community C(E)TR for a given patient had to be at least six months after the last, to ensure there was no overlap of outcome windows within the same patients.

### Supplementary Text S6. Age and sex demographics in general SLaM population of people with ID and/or autistic people

Control groups were initially matched to the C(E)TR groups based on age and sex, as a first step to reduce any case-control differences in hospitalization rates and other outcomes that may be explained by these variables. This meant, however, that our description of age and sex profiles may represent a C(E)TR cohort that is notably different to the rest of the SLaM population of patients with ID and/or autistic patients, or it may be truly representative of the populations utilizing these services in South London. We interpret our results in the context of an extraction on the full population of ID and autistic people utilizing SLaM services as of July 2025. Since analyses in the current study cover the years 2015 - 2025, this does not rule out changes in SLaM demographics across this period. It can, however, provide us with a recent picture of these demographics. Of 4273 patients with ID, 61.5% were male, with a median age of 27 years (28 IQR). There were 7557 autistic patients without ID, 64.7% of whom were male, with a median age of 20 years (12 IQR). Although the ID patients receiving a community C(E)TR align with these demographics, the autistic group of people receiving community C(E)TRs represents relatively more female patients who were also younger than the underlying SLaM population of autistic patients.

**Supplementary Figure S1.**
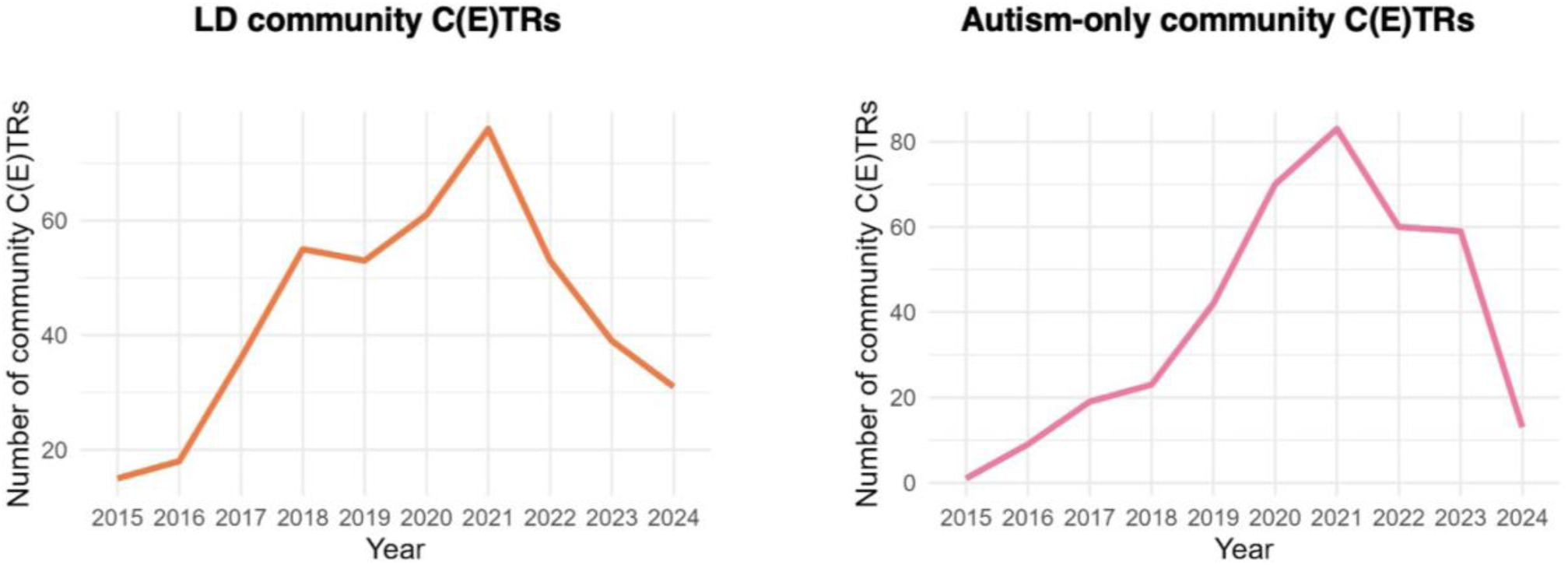
Community C(E)TR timeline. Number of identified community C(E)TRs per year at South London and Maudsley (SLaM) NHS Foundation Trust. Note that the y-axis in each graph is different.

**Supplementary Table S1.** Standardized mean difference (SMD) between case and control groups for the included covariates after inverse probability weighting for the primary outcome six-month analysis.

|  | **Mean Treated** | **Mean Untreated** | **Standardised Mean Diff (SMD)** |
|---|---|---|---|
| Age (mean years) | 25.93 | 24.92 | 0.08 |
| Sex (females) | 0.35 | 0.41 | -0.12 |
| Group (Autism only) | 0.36 | 0.38 | -0.04 |
| IMD Deprivation score (mean) | 25.04 | 25.78 | -0.06 |
| Substance use disorders (ICD10 F1*) | 0.05 | 0.06 | -0.03 |
| Schizophrenia spectrum disorders (ICD10 F2*) | 0.28 | 0.26 | 0.05 |
| Mood (affective) disorders (ICD10 F3*) | 0.12 | 0.12 | 0.00 |
| Anxiety disorders (ICD10 F4*) | 0.22 | 0.21 | 0.02 |
| Behavioural syndromes associated with physiological disturbances and physical factors (ICD10 F5*) | 0.02 | 0.02 | -0.03 |
| Personality disorder (ICD10 F6*) | 0.07 | 0.07 | 0.03 |
| Behavioural and emotional disorders with onset in childhood, incl. ADHD (ICD10 F9*) | 0.21 | 0.27 | -0.14 |
| Antipsychotic medication prescription | 0.66 | 0.68 | -0.05 |
| Antidepressant medication prescription | 0.63 | 0.63 | 0.01 |
| Mood stabiliser medication prescription | 0.36 | 0.37 | -0.00 |
| ADHD medication prescription | 0.24 | 0.28 | -0.09 |
| Hypnotic / anxiolytic medication prescription | 0.73 | 0.71 | 0.03 |
| Ethnicity |  |  |  |
| Asian | 0.05 | 0.07 | -0.08 |
| Black | 0.31 | 0.34 | -0.06 |
| Mixed Race | 0.09 | 0.09 | 0.01 |
| Other | 0.04 | 0.02 | 0.10 |
| White | 0.51 | 0.48 | 0.05 |

The efficacy of inverse probability weighting (IPTW) was evaluated by assessing the standardized mean difference (SMD) between case and control groups for the included covariates after IPTW. The table shows IPTW for one of the five imputations. Due to low levels of missing data (Ethnicity: 4.1%; Deprivation score: 1.3%), the five imputations showed very similar SMDs after IPTW. SMDs > -0.10 and < 0.10 are considered acceptable and unlikely to contribute to meaningful confounding. Most variables showed acceptable levels of SMD after IPW (all p>.05). The ‘Group’ variable represents the intellectual disability group (with or without autism) coded as 0, or the autism group (without intellectual disability) coded as 1. Sex was coded as 0 for male and 1 for female. The use of propensity weights in the difference-in-difference analysis following this step provides a doubly-robust framework for estimating differences in the primary outcome variable.

**Supplementary Table S2.**
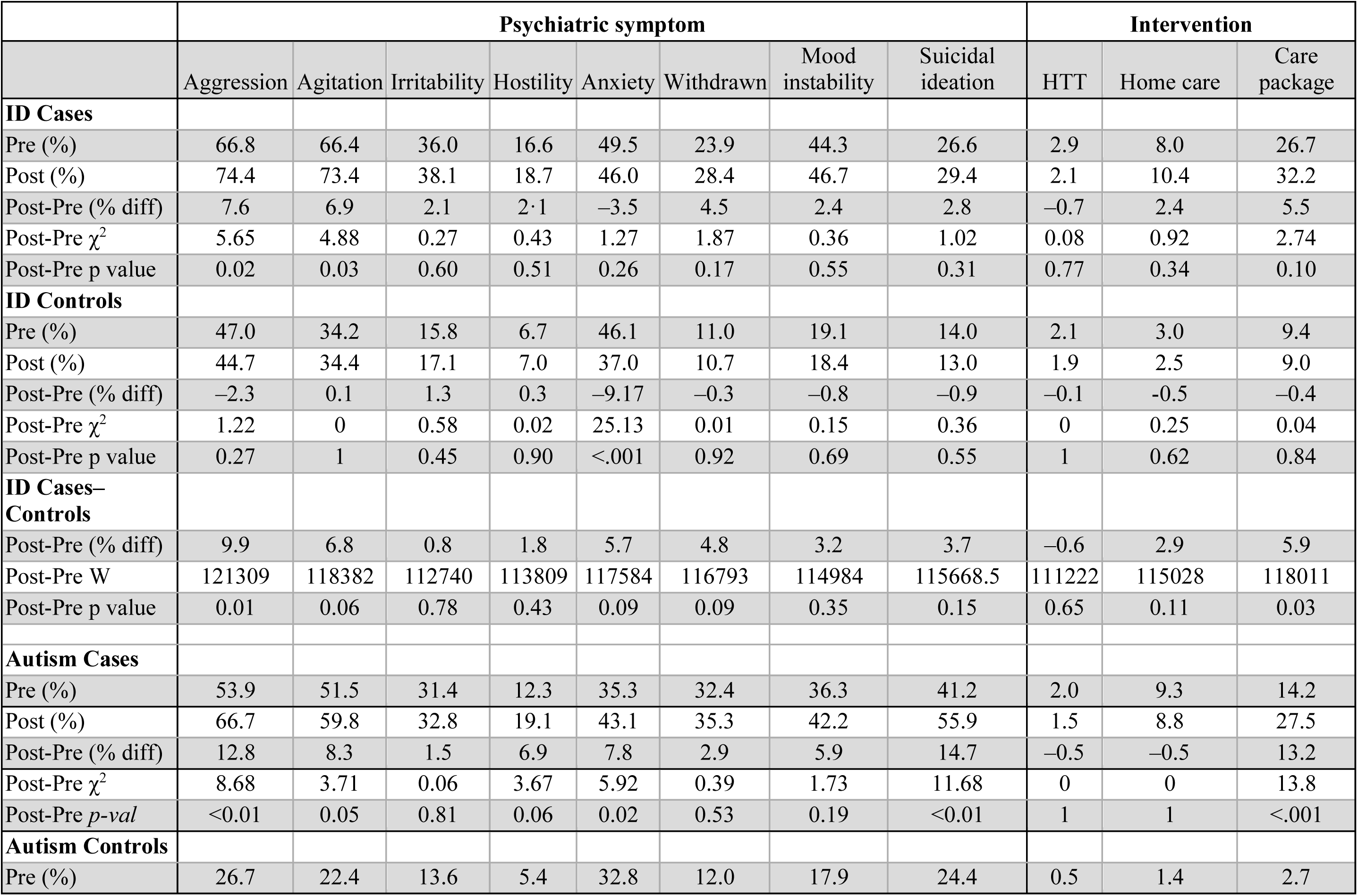
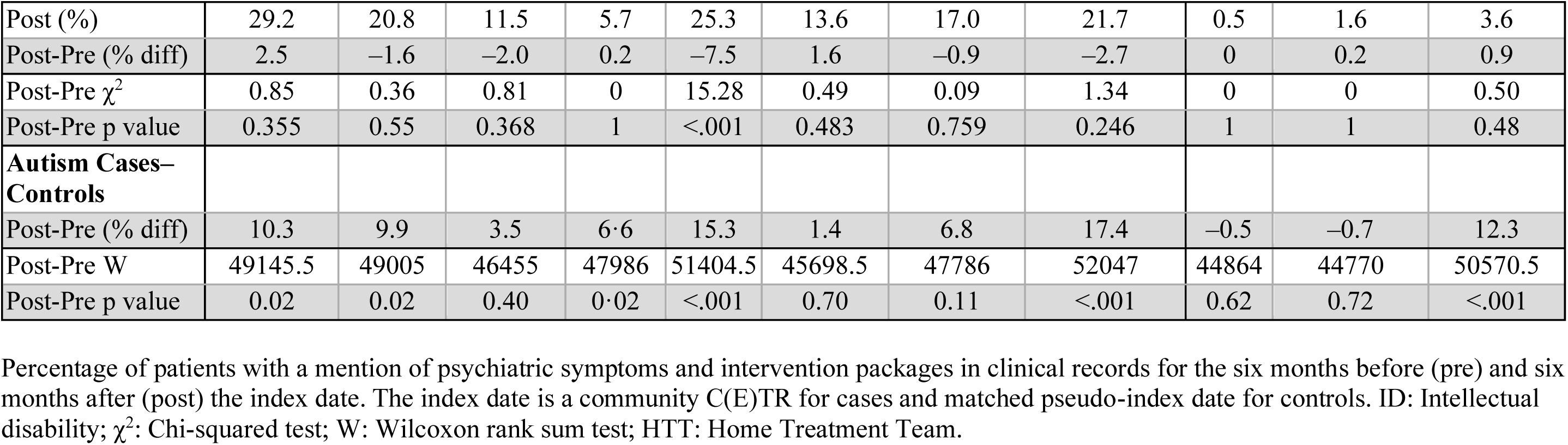
Mention of psychiatric symptoms and intervention packages in patients’ records.

|  | **Psychiatric symptom** |  |  |  |  |  |  |  | **Intervention** |  |  |
|---|---|---|---|---|---|---|---|---|---|---|---|
|  | Aggression | Agitation | Irritability | Hostility | Anxiety | Withdrawn | Mood instability | Suicidal ideation | HTT | Home care | Care package |
| **ID Cases** |  |  |  |  |  |  |  |  |  |  |  |
| Pre (%) | 66.8 | 66.4 | 36.0 | 16.6 | 49.5 | 23.9 | 44.3 | 26.6 | 2.9 | 8.0 | 26.7 |
| Post (%) | 74.4 | 73.4 | 38.1 | 18.7 | 46.0 | 28.4 | 46.7 | 29.4 | 2.1 | 10.4 | 32.2 |
| Post-Pre (% diff) | 7.6 | 6.9 | 2.1 | 2·1 | –3.5 | 4.5 | 2.4 | 2.8 | –0.7 | 2.4 | 5.5 |
| Post-Pre χ<sup>2</sup> | 5.65 | 4.88 | 0.27 | 0.43 | 1.27 | 1.87 | 0.36 | 1.02 | 0.08 | 0.92 | 2.74 |
| Post-Pre p value | 0.02 | 0.03 | 0.60 | 0.51 | 0.26 | 0.17 | 0.55 | 0.31 | 0.77 | 0.34 | 0.10 |
| **ID Controls** |  |  |  |  |  |  |  |  |  |  |  |
| Pre (%) | 47.0 | 34.2 | 15.8 | 6.7 | 46.1 | 11.0 | 19.1 | 14.0 | 2.1 | 3.0 | 9.4 |
| Post (%) | 44.7 | 34.4 | 17.1 | 7.0 | 37.0 | 10.7 | 18.4 | 13.0 | 1.9 | 2.5 | 9.0 |
| Post-Pre (% diff) | –2.3 | 0.1 | 1.3 | 0.3 | –9.17 | –0.3 | –0.8 | –0.9 | –0.1 | -0.5 | –0.4 |
| Post-Pre χ<sup>2</sup> | 1.22 | 0 | 0.58 | 0.02 | 25.13 | 0.01 | 0.15 | 0.36 | 0 | 0.25 | 0.04 |
| Post-Pre p value | 0.27 | 1 | 0.45 | 0.90 | <.001 | 0.92 | 0.69 | 0.55 | 1 | 0.62 | 0.84 |
| **ID Cases–Controls** |  |  |  |  |  |  |  |  |  |  |  |
| Post-Pre (% diff) | 9.9 | 6.8 | 0.8 | 1.8 | 5.7 | 4.8 | 3.2 | 3.7 | –0.6 | 2.9 | 5.9 |
| Post-Pre W | 121309 | 118382 | 112740 | 113809 | 117584 | 116793 | 114984 | 115668.5 | 111222 | 115028 | 118011 |
| Post-Pre p value | 0.01 | 0.06 | 0.78 | 0.43 | 0.09 | 0.09 | 0.35 | 0.15 | 0.65 | 0.11 | 0.03 |
| **Autism Cases** |  |  |  |  |  |  |  |  |  |  |  |
| Pre (%) | 53.9 | 51.5 | 31.4 | 12.3 | 35.3 | 32.4 | 36.3 | 41.2 | 2.0 | 9.3 | 14.2 |
| Post (%) | 66.7 | 59.8 | 32.8 | 19.1 | 43.1 | 35.3 | 42.2 | 55.9 | 1.5 | 8.8 | 27.5 |
| Post-Pre (% diff) | 12.8 | 8.3 | 1.5 | 6.9 | 7.8 | 2.9 | 5.9 | 14.7 | –0.5 | –0.5 | 13.2 |
| Post-Pre χ<sup>2</sup> | 8.68 | 3.71 | 0.06 | 3.67 | 5.92 | 0.39 | 1.73 | 11.68 | 0 | 0 | 13.8 |
| Post-Pre *p-val* | <0.01 | 0.05 | 0.81 | 0.06 | 0.02 | 0.53 | 0.19 | <0.01 | 1 | 1 | <.001 |
| **Autism Controls** |  |  |  |  |  |  |  |  |  |  |  |
| Pre (%) | 26.7 | 22.4 | 13.6 | 5.4 | 32.8 | 12.0 | 17.9 | 24.4 | 0.5 | 1.4 | 2.7 |
| Post (%) | 29.2 | 20.8 | 11.5 | 5.7 | 25.3 | 13.6 | 17.0 | 21.7 | 0.5 | 1.6 | 3.6 |
| Post-Pre (% diff) | 2.5 | –1.6 | –2.0 | 0.2 | –7.5 | 1.6 | –0.9 | –2.7 | 0 | 0.2 | 0.9 |
| Post-Pre χ<sup>2</sup> | 0.85 | 0.36 | 0.81 | 0 | 15.28 | 0.49 | 0.09 | 1.34 | 0 | 0 | 0.50 |
| Post-Pre p value | 0.355 | 0.55 | 0.368 | 1 | <.001 | 0.483 | 0.759 | 0.246 | 1 | 1 | 0.48 |
| **Autism Cases–Controls** |  |  |  |  |  |  |  |  |  |  |  |
| Post-Pre (% diff) | 10.3 | 9.9 | 3.5 | 6·6 | 15.3 | 1.4 | 6.8 | 17.4 | –0.5 | –0.7 | 12.3 |
| Post-Pre W | 49145.5 | 49005 | 46455 | 47986 | 51404.5 | 45698.5 | 47786 | 52047 | 44864 | 44770 | 50570.5 |
| Post-Pre p value | 0.02 | 0.02 | 0.40 | 0·02 | <.001 | 0.70 | 0.11 | <.001 | 0.62 | 0.72 | <.001 |

Percentage of patients with a mention of psychiatric symptoms and intervention packages in clinical records for the six months before (pre) and six months after (post) the index date. The index date is a community C(E)TR for cases and matched pseudo-index date for controls. ID: Intellectual disability; χ<sup>2</sup>: Chi-squared test; W: Wilcoxon rank sum test; HTT: Home Treatment Team.

**Supplementary Table S3.** Direct community contacts with healthcare professionals.

|  | Type of Professional |  |  |  |  |  |  |  |  |
|---|---|---|---|---|---|---|---|---|---|
|  | Clinical Support Worker | Nurse | OT | Pharmacist | Physiotherapist | Psychiatrist | Psychologist / Therapist | Social Worker | SLT |
| **ID Cases** |  |  |  |  |  |  |  |  |  |
| Pre # (SD) | 0.17 (1.33) | 8.50 (13.34) | 0.72 (2.77) | 0.07 (0.37) | 0.02 (0.20) | 4.91 (14.94) | 3.80 (10.43) | 0.93 (2.12) | 0.02 (0.26) |
| Post # (SD) | 0.18 (1.10) | 8.03 (13.89) | 0.62 (3.58) | 0.23 (1.60) | 0.01 (0.13) | 3.09 (6.53) | 4.04 (10.79) | 0.88 (2.55) | 0.00 (0.06) |
| Post-Pre (#) | 0.00 | -0.47 | -0.10 | 0.15 | -0.01 | -1.82 | 0.24 | -0.05 | -0.01 |
| Post-Pre *V* | 90 | 8805 | 197.5 | 90.5 | 2 | 4478.5 | 2598.5 | 2309 | 1 |
| Post-Pre *p-val* | 0.86 | 0.55 | 0.22 | 0.52 | 0.79 | 0.34 | 0.43 | 0.68 | 1 |
| **ID Controls** |  |  |  |  |  |  |  |  |  |
| Pre (#) | 0.44 (1.98) | 10.48 (15.86) | 0.57 (2.29) | 0.57 (2.28) | 0.00 (0.06) | 2.94 (4.41) | 2.19 (6.45) | 0.94 (4.13) | 0.01 (0.09) |
| Post (#) | 0.45 (1.96) | 10.53 (16.16) | 0.56 (2.49) | 0.72 (2.57) | 0.02 (0.19) | 2.82 (3.88) | 1.98 (5.92) | 0.86 (3.23) | 0.01 (0.15) |
| Post-Pre (#) | 0.00 | 0.05 | -0.02 | 0.15 | 0.01 | -0.13 | -0.21 | -0.08 | 0.01 |
| Post-Pre *V* | 1449 | 30076 | 2139 | 3077 | 29 | 51684 | 12398 | 5878.5 | 10 |
| Post-Pre *p-val* | 0.65 | 0.67 | 0.6 | 0.08 | 0.12 | 0.48 | 0.66 | 0.79 | 0.59 |
| **ID Cases-Controls** |  |  |  |  |  |  |  |  |  |
| Post-Pre *V* | 71226 | 69248 | 68928 | 68900 | 69975 | 70129 | 71743 | 70541 | 70623 |
| Post-Pre *p-val* | 0.78 | 0.63 | 0.36 | 0.33 | 0.22 | 0.84 | 0.72 | 0.94 | 0.81 |
| **Autism Cases** |  |  |  |  |  |  |  |  |  |
| Pre (#) | 1.90 (8.81) | 6.95 (16.55) | 3.86 (16.06) | 0.05 (0.22) | 0.00 (0.00) | 3.21 (5.08) | 3.29 (9.26) | 1.18 (4.07) | 0.30 (2.44) |
| Post (#) | 1.43 (5.79) | 6.07 (10.58) | 2.52 (11.42) | 0.12 (0.43) | 0.07 (0.60) | 3.91 (7.50) | 2.31 (6.31) | 0.86 (2.42) | 0.30 (3.07) |
| Post-Pre (#) | -0.47 | -0.88 | -1.34 | 0.07 | 0.07 | 0.71 | -0.98 | -0.31 | 0.00 |
| Post-Pre *V* | 87 | 995.5 | 191 | 49.5 | 3 | 1304 | 329 | 312.5 | 3 |
| Post-Pre *p-val* | 0.97 | 0.72 | 0.27 | 0.12 | 0.37 | 0.96 | 0.74 | 0.4 | 1 |
| **Autism Controls** |  |  |  |  |  |  |  |  |  |
| Pre (#) | 0.67 (2.87) | 6.74 (15.20) | 1.41 (5.00) | 0.07 (0.45) | 0.09 (1.06) | 2.44 (4.89) | 2.74 (9.59) | 0.55 (1.93) | 0.01 (0.11) |
| Post (#) | 0.58 (2.47) | 7.74 (19.57) | 0.89 (3.34) | 0.11 (0.84) | 0.04 (0.34) | 2.28 (5.30) | 2.96 (15.68) | 0.85 (3.55) | 0.00 (0.00) |
| Post-Pre (#) | -0.09 | 1.01 | -0.52 | 0.04 | -0.05 | -0.15 | 0.22 | 0.30 | -0.01 |
| Post-Pre *V* | 184.5 | 2124 | 169 | 27.5 | 1 | 1938.5 | 686 | 300.5 | 0 |
| Post-Pre *p-val* | 0.83 | 0.82 | 0.19 | 0.59 | 1 | 0.54 | 0.36 | 0.3 | 0.35 |
| **Autism Cases-Controls** |  |  |  |  |  |  |  |  |  |
| Post-Pre *V* | 7743.5 | 8130.5 | 7720.5 | 7920.5 | 7851 | 7743 | 7878.5 | 6828.5 | 7725.5 |
| Post-Pre *p-val* | 0.91 | 0.44 | 0.97 | 0.41 | 0.23 | 0.94 | 0.72 | 0.05 | 0.87 |

Direct community contacts with clinical professionals in the six months before and after a community C(E)TR (cases) / matched index dates (controls). The number (#, SD) of contacts are shown and the differences per group and between groups is captured using a Wilcoxin signed rank test (*V*). ID = Intellectual Disability; OT = Occupational Therapist; SLT = Speech and Language Therapist.

## References

1. Hossain MM, Khan N, Sultana A, Ma P, McKyer ELJ, Ahmed HU, Purohit N. Prevalence of comorbid psychiatric disorders among people with autism spectrum disorder: An umbrella review of systematic reviews and meta-analyses. Psychiatry Res 2020; 287:112922. doi: 10.1016/j.psychres.2020.112922. Epub 2020 Mar 18. PMID: 32203749.

2. Mazza MG, Rossetti A, Crespi G, Clerici M. Prevalence of co-occurring psychiatric disorders in adults and adolescents with intellectual disability: A systematic review and meta-analysis. J Appl Res Intellect Disabil 2020; 33(2):126–138. doi: 10.1111/jar.12654.

3. Bowring DL, Totsika V, Hastings RP, Toogood S, Griffith GM. Challenging behaviours in adults with an intellectual disability: a total population study and exploration of risk indices. Brit J Clin Psychol 2017; 56: 16–32

4. Rattaz C, Michelon C, Munir K, Baghdadli A. Challenging behaviours at early adulthood in autism spectrum disorders: topography, risk factors and evolution. J Intell Dis Res 2018; 62(7): 637–649.

5. NHS England. Building the right support: A national plan to develop community services and close inpatient facilities for people with a learning disability and/or autism who display behaviour that challenges, including those with a mental health condition. https://www.england.nhs.uk/wp-content/uploads/2015/10/ld-nat-imp-plan-oct15.pdf, Date: Dec 02, 2015, Date accessed: Oct 07, 2026

6. NHS England. Supporting people with learning disability and/or autism who have a mental health condition or display behaviour that challenges. https://www.england.nhs.uk/wp-content/uploads/2015/10/service-model-291015.pdf, Date: Oct, 2015, Date accessed: Oct 07, 2026

7. NHS England. Transforming care—model service specifications: Supporting implementation of the service model. https://www.england.nhs.uk/publication/transforming-care-service-model-specification-january-2017/, Date: Feb 03, 2017, Date accessed: Oct 07, 2026

8. Lindgren S, Lauer E, Momany E, et al. Disability, Hospital Care, and Cost: Utilization of Emergency and Inpatient Care by a Cohort of Children with Intellectual and Developmental Disabilities. J Pediatrics 2021; 229: 259–266. doi: 10.1016/j.jpeds.2020.08.084.

9. Nieuwenhuis JG, Lepping P, Mulder NL, Nijman HLI, Veereschild M, Noorthoorn EO. Increased prevalence of intellectual disabilities in higher-intensity mental healthcare settings. BJPsych Open 2021; 7(3): e83. doi:10.1192/bjo.2021.28

10. Henderson A, Mcskimming P, Kinnear D, et al. Changes over a decade in psychotropic prescribing for people with intellectual disabilities: prospective cohort study. BMJ Open 2020; 10: e036862. doi: 10.1136/bmjopen-2020-036862

11. Jones LJ, Woodrow C. Adherence to care and treatment review policy and impact on length of stay. Adv Ment Health Intell Disabil 2023; 17(4): 283–291. doi: 10.1108/AMHID-07-2023-0023

12. Langdon PE, Thompson PA, Shepstone L, et al. Psychiatric inpatient admissions and discharges of people with intellectual disabilities: A time series analysis of English national data. J Appl Res Intellect Disabil 2023; 36(1):196–204. doi: 10.1111/jar.13051

13. McCoy B, Bell L, Wang K, et al. Investigating the use and impact of community Care (Education) and Treatment Reviews (C(E)TRs) in people with intellectual disability and autistic people: protocol for a cohort study using electronic health records. BMJ Open 2025; 15: e107889. doi:10.1136/ bmjopen-2025-107889

14. NIHR Maudsley Biomedical Research Centre Clinical Record Interactive Search (CRIS): Natural language processing service. https://www.maudsleybrc.nihr.ac.uk/facilities/clinical-record-interactive-search-cris/natural-language-processing-nlp-service/, Date accessed: 07 Oct 2026.

15. Wing JK, Beevor AS, Curtis RH, Park SB, Hadden S, Burns A. Health of the Nation Outcome Scales (HoNOS). Research and development. Brit J Psychiatry 1998; 172: 11–8

16. Austin PC. An introduction to propensity score methods for reducing the effects of confounding in observational studies. Multivariate Behav Res 2011; 46: 399–424.

17. Stuart EA. Matching methods for causal inference: A review and a look forward. Stat Sci 2010; 25(1):1–21. doi: 10.1214/09-STS313. PMID: 20871802; PMCID: PMC2943670.

18. Stuart EA, Huskamp HA, Duckworth K, et al. Using propensity scores in difference-in-differences models to estimate the effects of a policy change. Health Serv Out Res Methodol 2014; 14:166–82.

19. Azur MJ, Stuart EA, Frangakis C. Leaf PJ. Multiple imputation by chained equations: what is it and how does it work? Int J Meth Psychiat Res 2011; 20: 40–49. 10.1002/mpr.329

20. Rubin DB. Multiple imputation for nonresponse in surveys. New York: John Wiley & Sons; 1987.

21. Knowles G, Gayer-Anderson C, Beards S, et al. Mental distress among young people in inner cities: the Resilience, Ethnicity and AdolesCent Mental Health (REACH) study. J Epidemiol Comm Health 2021; 75: 515–522.

22. Stewart G, Happé F. Aging Across the Autism Spectrum. Ann Rev Dev Psychol 2025; 7: 461–484. 10.1146/annurev-devpsych-111323-090813

23. NHS England. Minimum standards for keyworking services for autistic children and young people and children and young people with a learning disability. https://www.england.nhs.uk/long-read/minimum-standards-for-keyworking-services-for-autistic-children-and-young-people-and-children-and-young-people-with-a-learning-disability/, Date: April 14, 2026, Date accessed: Oct 07, 2026

24. Abraham J, Purandare K, McCabe J, et al. An 8-year study of admissions and discharges to a specialist intellectual disability inpatient unit. J Appl Res Intellect Disabil 2022; 35(2):569–576. doi: 10.1111/jar.12967

25. NHS Digital. Statistics on people with a learning disability and autistic people in mental health hospitals. https://digital.nhs.uk/data-and-information/publications/statistical/learning-disability-services-statistics, Date accessed: Oct 07, 2026

26. NHS England. Care and Treatment review: Policy and Guidance. https://www.england.nhs.uk/wp-content/uploads/2023/01/Dynamic-support-register-and-Care-Education-and-Treatment-Review-policy-and-guide.pdf, Date: Jan, 2023, Date accessed: 07 Oct 2026

27. Amiola A, Patteril E, Chester V, et al. Perceptions to Care, (Education) and Treatment Reviews (C(E)TRs) of Mental Health Clinicians Working with Adults with Intellectual Disability in England: A Cross-Sectional Study. J Men Health Res Intell Disabil 2026; 19(1): 43–75. 10.1080/19315864.2025.2525834

28. NHS England. Community mental health framework for adults and older adults. https://www.england.nhs.uk/publication/the-community-mental-health-framework-for-adults-and-older-adults/, Date: Sept 29, 2019, Date accessed: Sept 7, 2026

29. Sheehan R, Mutch J, Marston L, Osborn D, Hassiotis A. Risk factors for in-patient admission among adults with intellectual disability and autism: investigation of electronic clinical records. BJPsych Open 2020; 7(1): e5. doi: 10.1192/bjo.2020.135.

30. Bohen F, Woodrow C. Dynamic support database clinical support tool: inter-rater reliability. Adv Ment Health Intell Disabil 2020; 14(2): 25–32

## References

1. NIHR Maudsley Biomedical Research Centre Clinical Record Interactive Search (CRIS): Natural language processing service. https://www.maudsleybrc.nihr.ac.uk/facilities/clinical-record-interactive-search-cris/natural-language-processing-nlp-service/ Date accessed: 07 Oct 2026.

2. Abadie A. Semiparametric Difference-in-Differences Estimators. Rev Econ Stud 2005; 72:1–19

3. Szmaragd C, Clarke P, Steele F.Subject specific and population average models for binary longitudinal data: a tutorial. Longitud life course stud 2013; 42(2): 147–165.

4. Heckmann J, Ichimura H, Todd PE. Matching as an Econometric Evaluation Estimator: Evidence from Evaluating a Job Training Programme. Rew Econ Stud 1997; 64:505–654

5. Stuart EA, Huskamp HA, Duckworth K, et al. Using propensity scores in difference-in-differences models to estimate the effects of a policy change. Health Serv Out Res Methodol 2014; 14:166–82.

